# Localising the epileptogenic zone from single-pulse electrical stimulation responses using cross-trial attention

**DOI:** 10.64898/2026.07.27.26358819

**Authors:** Jamie Norris, Dorien van Blooijs, Aswin Chari, Gerald Cooray, Martin Tisdall, Karl Friston, Stuart D W Smith, Richard Rosch

**Author notes:** **Corresponding author:** Jamie Norris, UCL Institute of Health Informatics, London, UK.

## Abstract

**Background:** Analysis of SPES responses often relies on averaging repeated stimulation trials to improve signal quality. However, this may obscure clinically relevant trial-to-trial variation. We tested whether explicitly modelling cross-trial dependencies improves localisation of the epileptogenic zone, using concordance with the clinical SOZ as a proxy endpoint, and explored whether resection of model-positive channels is associated with postsurgical seizure freedom.

**Methods:** We developed an interleaved Hierarchical Attention Transformer (HAT), a deep learning architecture that models cross-trial and cross-channel dependencies in SPES responses without averaging. We compared it with two baselines averaging responses or trial embeddings. Models were evaluated with patient-held-out, repeated five-fold cross-validation on SPES data from 35 patients. Robustness to reduced trial availability at inference was assessed by restricting test inputs to 1 or 5 trials. Associations with surgical outcome were assessed using AUROC and patient-level tests on the proportion of model-positive channels resected.

**Results:** The HAT had higher SOZ concordance than the trial-averaged baseline (AUROC 0.762 vs 0.721; mean paired difference 0.041; one-sided 95% lower confidence bound 0.009; Holm-adjusted p = 0.0197). Performance changed little when inference was restricted to 1 trial. Outcome analyses did not provide statistical evidence that seizure-free patients had a higher proportion of model-positive channels resected (AUROC 0.634; p = 0.102).

**Conclusions:** Modelling cross-trial dependencies improved concordance with the SOZ compared with trial-averaged approaches, while remaining robust to reduced trial availability at inference. Associations with postsurgical outcome were inconclusive, consistent with limited sample size and training on SOZ labels rather than outcome-aligned labels.

**Key points:**

- Cross-trial modelling of SPES responses improved concordance with the clinical SOZ compared with trial-averaged baselines.
- In a 35-patient cohort, SOZ AUROC was higher than a trial-averaged baseline (0.762 vs 0.721).
- Performance was stable when inference used as few as 1 trial, with the largest drop in the trial-averaged baseline.
- Resection of model-positive channels did not provide statistical evidence of an association with seizure freedom (AUROC 0.634; one-sided Mann–Whitney p = 0.102).

## Introduction

Epilepsy disrupts normal brain activity, leading to seizures and abnormal brain discharges between seizures, both of which arise from dynamic, patient-specific networks. For the approximately 7 in 1000 people diagnosed with epilepsy^1^, achieving optimal seizure control often requires highly individualised medical treatments. Notably, around one-third of cases are resistant to medication, posing significant challenges for effective seizure management^2^. In these drug-resistant cases, surgery can be a viable treatment option, with the aim of reducing or eliminating seizures by resecting or disconnecting brain regions responsible for seizure generation, collectively referred to as the epileptogenic zone. The best current approach to delineate a putative epileptogenic zone relies on mapping brain areas involved in seizure onset^3^. Accurate identification of the seizure onset zone (SOZ) is essential to achieve favourable surgical outcomes, ideally leading to postoperative seizure freedom. Non-invasive techniques like scalp EEG and neuroimaging provide useful information about putative seizure onset zones, but in complex cases, more precise identification often requires invasive intracranial EEG recordings. Intracranial recording places a considerable burden on patients, often requiring several days of monitoring to capture a sufficient number of habitual seizures for reliable delineation of the SOZ. Moreover, for around 15% of patients, no clear SOZ is identified, preventing surgery^4^. Even when surgery is performed, only about half of patients achieve seizure freedom at a five-year follow-up^5^. Complete resection of the clinically defined SOZ does not guarantee seizure freedom, and the relationship between the proportion of SOZ contacts resected and outcome remains unclear^6–8^. This uncertainty highlights the limitations of relying solely on the passively defined SOZ and the need for complementary approaches to characterise epileptogenic tissue.

In addition to passive monitoring of spontaneous seizures, single-pulse electrical stimulation (SPES) allows clinicians to actively probe the epileptic brain (**Figure 1A**). During SPES, isolated electrical stimuli are applied to pairs of intracranial electrodes, and the resulting responses are recorded on contacts within the cortex of other brain regions^9^. These responses reflect the connectivity between stimulated and recorded areas, allowing for the mapping of directed (effective) connectivity and the assessment of excitability within implanted brain regions. Typically, targeted regions are stimulated 10 to 50 times, and the responses are averaged across trials to improve the signal-to-noise ratio.

**Figure 1.**
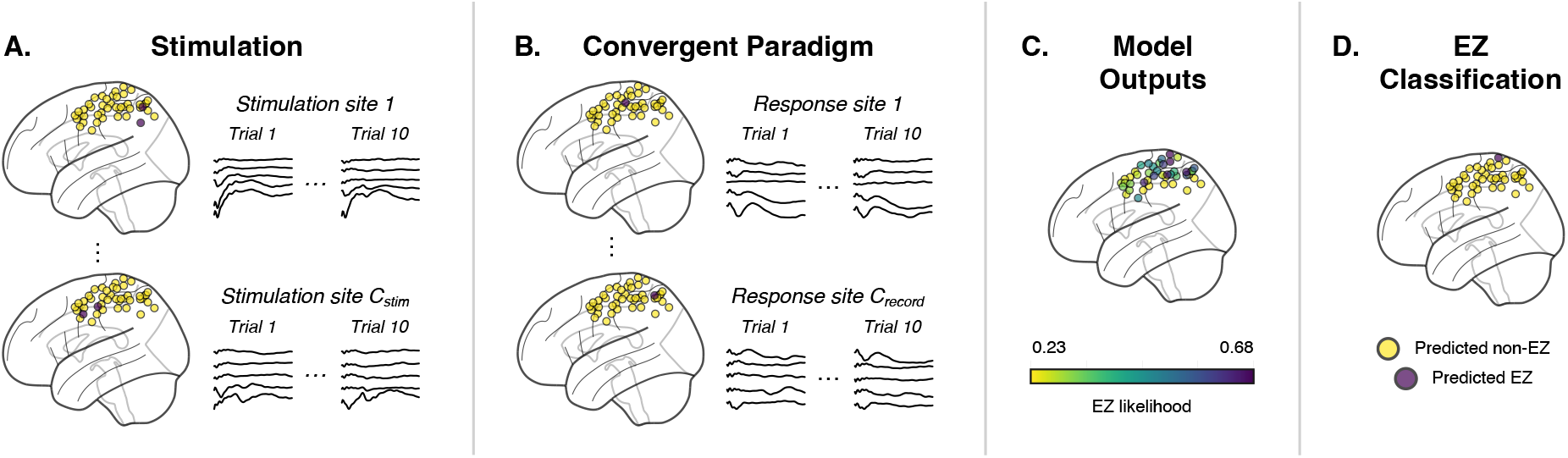
EZ localisation pipeline for an example patient. (**A)** Stimulation is performed at pairs of adjacent channels. (**Top**) Each stimulation train consists of multiple SPES trials, with stimulation applied at the site shown in purple and responses recorded at different sites (yellow). Each row corresponds to a unique response site. (**Bottom**) The same process is shown for a different stimulation site. **(B)** Responses are aggregated using a convergent paradigm, where responses are grouped by recording site rather than stimulation site. (**Top**) iEEG traces for a single response site (purple) are shown, with each row recorded at the same channel during stimulation at different sites (yellow). Trials 1 and 10 are displayed. (**Bottom**) Traces for a second response site are shown. **(C)** Models are trained using the clinical SOZ labels and produce a score from 0 to 1 for each response channel. **(D)** A patient-specific threshold is applied, with scores at or above the threshold designated model positive.

Various machine learning approaches have been used to automatically identify the seizure onset zone from SPES responses^10–13^. Initial studies trained models on individual responses from single channels, treating each response independently^10,11^, with subsequent work utilising multi-channel responses with a convolutional neural network^12^. More recently, Transformer models with cross-channel attention have been applied^13^, enabling the use of inputs with variable electrode numbers and placements, as is typical in intracranial EEG. While these methods show promising performance, they either classify individual trials, resulting in multiple predictions per site, or take trial-averaged responses as input, which may obscure clinically meaningful features. For example, delayed responses, which occur in only a subset of trials and are not time-locked to the stimulus, have been linked to epileptogenicity^14^, and the removal of regions exhibiting such responses has been associated with positive surgical outcomes^15^. Similarly, trial-by-trial response variability has been linked to epileptogenicity^16,17^, further highlighting the potential limitations of relying solely on trial-averaged data. Incorporating multi-trial responses directly, without the need to average, may preserve clinically relevant information.

We introduce an interleaved Hierarchical Attention Transformer (HAT) that models cross-trial and cross-channel dependencies in multi-trial SPES without prior averaging. We evaluate whether modelling cross-trial dependencies improves SOZ concordance relative to trial-averaged baselines, quantify how performance changes when the number of trials available at inference is reduced, and test whether overlap between resected tissue and model-positive channels is associated with postsurgical outcome. The HAT increased AUROC by 0.041 over the trial-averaged baseline, but the exploratory outcome analysis did not demonstrate clinical utility.

## Materials and methods

### Study design and scope

We used patient-held-out, repeated five-fold cross-validation on a single-centre SPES cohort. The following sections describe the dataset, preprocessing, model architectures, training procedures, and evaluation. This was a retrospective secondary analysis. All patients in the released dataset with the channel-level labels required for this analysis were included; no prospective sample-size calculation was performed and no participant was excluded on the basis of model performance.

### Study participants and data collection

The open-source dataset used in this study originates from a clinical study conducted at the University Medical Center Utrecht, the Netherlands^18^. It comprises electrocorticography recordings from 74 patients who underwent clinically indicated single-pulse electrical stimulation between 2012 and 2020. Of these, 35 patients had available seizure onset zone labels and were included in the present study. The source cohort excluded patients with large structural lesions and required electrode positions to be localisable from computed tomography co-registered to T1-weighted magnetic resonance imaging.

The analysed cohort had a mean age of 21.9 years (standard deviation 13.0), a median age of 17 years (interquartile range 13.5–31.5; range 4–51), and included 16 female and 19 male patients. Eighteen participants were younger than 18 years and 17 were adults. Children and adults were analysed together, and age-stratified performance was not estimated. A median of 54 bipolar stimulation sites were sampled per patient (interquartile range 47.5–58.5), with approximately 10 trials per stimulation site, while the median number of SOZ contacts was 8 (interquartile range 4.5–12).

Across the 35 participants, the public release contained 2,474 implanted contacts, comprising 1,896 grid, 448 strip and 130 depth contacts. The median number of contacts per patient was 64 (range 48–112). Implantation was left unilateral in 22 patients, right unilateral in 12 and bilateral in one. Recording channels, including those within the SOZ, were sampled across all major cortical lobes. Postsurgical seizure outcome was assessed at five years: 12 patients were seizure-free and 23 were not. Patient-level cohort and stimulation details are provided in Supporting Methods S1 and Supplementary Tables S1 and S2.

### Ethics and consent

The source study was approved by the Medical Research Ethics Committee of University Medical Center Utrecht and complied with the relevant ethical regulations. Informed consent was waived for patients included from January 2008 to December 2017; explicit consent for research use was obtained from January 2018 onwards. The released data were anonymised and the present study involved no further participant contact.

### Recording and stimulation protocol

ECoG was recorded at 2048 Hz using subdural grids and strips with a 4.2 mm² contact surface and 10 mm inter-contact spacing. SPES comprised ten 1 ms monophasic pulses delivered at 0.2 Hz through adjacent contacts in a bipolar configuration. Current was normally 8 mA and was reduced to 4 mA near central nerves or primary sensorimotor cortex; 94.8% of stimulation events in the analysed cohort used 8 mA and 5.2% used 4 mA.

### Preprocessing

Preprocessing was adapted from Norris *et al.*^13^ and van Blooijs *et al*^19^. Signals were band-pass filtered (1–150 Hz) and notch-filtered at 50 Hz and its harmonics to remove alternating current mains electrical interference. The dataset includes annotations for artefacts and seizures, and all trials overlapping these events were excluded. To mitigate stimulation artefacts propagating to nearby electrodes through volume conduction, we followed van Blooijs *et al.*^19^, excluding the first 9 milliseconds post-stimulation and any channels within 13 mm of the stimulation site. The subsequent 1-second period of each trial was used and downsampled to 512 Hz. Filtering was applied before cropping, and no additional offline re-referencing was performed beyond the released montage.

The data were organised under a convergent paradigm, a method previously shown to improve classification performance on the same dataset^13^. Unlike the divergent approach, which analyses responses at other sites following stimulation at a specific location, the convergent paradigm focuses on how a particular site responds to stimulation from various other locations (**Figure 1B**). This approach captures the input to each site and therefore provides a measure of its inherent excitability.

For each response site, the data were structured as *C_stim_* × *T* sets of trials, where *C_stim_* represents the number of stimulation sites and *T* is the number of trials.

### Models

The model architectures are summarised in **Figure 2**. A Transformer uses self-attention to learn how strongly each input should influence the others. Here, each token is a compact numerical representation of a SPES response. A learnable classification ([CLS]) token gathers information from the input tokens, and a linear classification head converts the resulting representation into one continuous channel score. Each model is adapted from Norris *et al.*^13^, with two key modifications. Firstly, the original convolutional neural network (CNN) was replaced with a multilayer perceptron (MLP) to reduce training time and enable a more thorough hyperparameter search. Secondly, in each model, the Euclidean distance between each stimulation-response site pair is concatenated to the MLP embedding to provide additional context.

**Figure 2.**
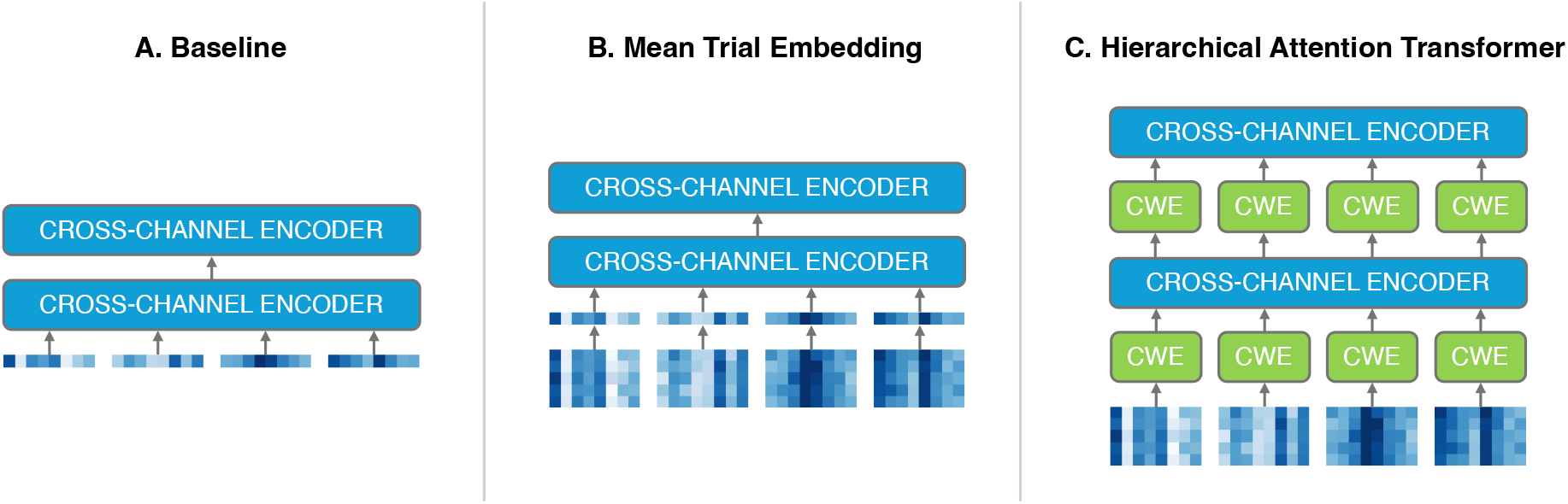
Model architectures. (**A) Baseline**: Responses are averaged across trials, embedded with an MLP, and input as tokens to the cross-channel encoder. **(B) Mean Trial Embedding (MTE)**: Each trial is embedded with the MLP, and mean pooling is applied across trial embeddings, which are then processed in the same manner as the baseline. **(C) Hierarchical Attention Transformer (HAT)**: Each trial is embedded with the MLP, and a channel-wise encoder (CWE) processes all trials per stimulation site. Each resulting stimulation-site representation serves as a token into the cross-channel encoder. The HAT consists of pairs of channel-wise and cross-channel layers. Blue boxes denote cross-channel encoders. Figure adapted from Chalkidis et al.^20^.

All models use an encoder-only Transformer with cross-channel self-attention. The resulting scores are continuous per-channel outputs trained to discriminate clinically identified SOZ from non-SOZ sites (**Figure 1C**); when binary labels are required for downstream analyses, patient-specific thresholds are applied to define a model-derived set of positive channels (**Figure 1D**).

#### Baseline

For the **Baseline** model, the trial-averaged SPES response for each stimulation site is embedded with the MLP, and its Euclidean distance to the recording site is concatenated to this embedding. The resulting tokens, together with a [CLS] token, are passed to the cross-channel Transformer.

#### Mean Trial Embedding (MTE)

The **MTE** model builds on the **Baseline** by embedding each SPES trial individually rather than averaging responses prior to embedding. The MLP processes each trial; the resulting embeddings are averaged across trials to form one token per stimulation site. These tokens (plus [CLS]) are then passed to the cross-channel Transformer.

#### Hierarchical Attention Transformer (HAT)

The **HAT** model extends the previous approaches by adding channel-wise (cross-trial) attention layers that are interleaved with the existing cross-channel layers, following the interleaved HAT variant proposed by Wu *et al*.^21^ and later extended by Chalkidis *et al.*^20^. Cross-trial attention captures relationships between repeated responses from the same stimulation-response site pair, whereas cross-channel attention captures relationships between responses at the same recording site following stimulation at different locations. SPES responses from all trials and channels are individually embedded by the MLP. A channel-wise (cross-trial) encoder is applied across trials for each stimulation site to capture cross-trial information; its outputs form site-level tokens, which then pass to a cross-channel encoder to model relationships across the stimulation sites contributing to the prediction for that recording site. We stack these channel-wise and cross-channel blocks in alternating pairs, enabling simultaneous modelling of cross-trial and cross-channel dependencies.

### Model Training

To evaluate the within-centre generalisability of each model, we applied repeated five-fold cross-validation with five repeats. For each repeat, patients were randomly assigned to five folds of seven, with a new assignment used for each repeat. Within each repeat, five iterations were run. In each iteration, all recordings from the 21 patients in three folds were pooled into one training set, one fold of seven patients was used for validation, and the remaining fold of seven patients was held out for testing. The validation and test roles rotated so that every fold served once as the test fold, producing five runs per repeat and 25 runs in total. Global means and standard deviations were computed on this training set for (1) all time-series data points and (2) all Euclidean distance values. These values were then used to standardise the training, validation, and test sets. During each training run, the area under the receiver operating characteristic curve (AUROC) was calculated for the validation set at each epoch. Early stopping was applied based on this validation AUROC, with a patience of 5 epochs (i.e. training stopped if the validation AUROC did not improve for five consecutive epochs) and a maximum of 25 epochs per training run. An epoch comprised one complete pass through the sampled training data. The repeated partitioning is described in Supporting Methods S2 and illustrated in Supplementary Figure S1.

Hyperparameter tuning was performed using the *Optuna* framework^22^, which automatically searched the hyperparameter space to maximise validation AUROC. During training, weighted sampling gave greater representation to SOZ sites and to patients with fewer candidate sites. A random proportion of valid trial-by-stimulation-site observations was also masked during each forward pass. Full details are provided in Supporting Methods S6. The held-out test fold was not used for standardisation, hyperparameter optimisation, early stopping, data augmentation, or threshold derivation. It was evaluated only after these procedures had been completed. All recordings from a given patient remained within the same training, validation, or test partition throughout each cross-validation split. All models were implemented in PyTorch^23^ and trained on a personal computer (Apple M1 Pro, 16 GB memory). Further details on hyperparameter search, sampling and augmentation, and training times are provided in Supporting Methods S5–S7 and Supplementary Tables S3–S6.

### Statistics and Reproducibility

#### SOZ concordance

We evaluated concordance with the clinically defined SOZ using AUROC, specificity, sensitivity, and Youden’s index (sensitivity + specificity – 1). Specificity, sensitivity, and Youden’s index were computed after applying patient-specific thresholds to the continuous model outputs. The threshold was derived without using the clinically defined SOZ labels of the test patient.

Thresholds that maximised Youden’s index in the validation patients were expressed relative to each patient’s median score and median absolute deviation (MAD), and the median scaling factor was then applied to the test patient. Channels scoring at or above the threshold were classified as model positive, and channels scoring below the threshold were classified as model negative. For example, if the median scaling factor was 2, a test patient median score of 0.20 and MAD of 0.05 would give a threshold of 0.30, meaning channels scoring at or above 0.30 would be classified as model positive. The full thresholding procedure is described in Supporting Methods S3.

For each metric, we report the mean across patients. Each patient contributed one held-out score from each of the five repeated cross-validation repetitions, and these five scores were averaged to give one value per patient before statistical inference. The two planned model comparisons were HAT versus Baseline and HAT versus MTE. Comparisons were paired because the same 35 patients were evaluated using each model. Mean AUROC differences were reported with one-sided 95% lower confidence bounds obtained from 10,000 patient-level bootstrap resamples. Statistical evidence for improvement was assessed using one-sided paired sign-flip permutation tests. These tests were used because the comparisons were paired and normality of the differences in AUROC between models was not assumed. The two p-values were adjusted using the Holm procedure. Paired Cohen’s d_z_ was calculated as the mean difference within patients divided by the standard deviation of those differences. Standard errors are reported to quantify uncertainty in the mean estimates. Details of the bootstrap, permutation test and standard error calculations are provided in Supporting Methods S4.

#### Trial Subset Analysis

We assessed the impact of reducing the number of trials available at inference. For each previously trained model, we restricted the test set to the first *n* trials (*n* = 1, 2, 5) and compared performance with the results obtained using all available trials (*n* ≤ 20).

#### Association of Model Predictions with Surgical Outcome

Finally, in an exploratory analysis of the relationship between model outputs and epileptogenicity, we investigated whether the proportion of channels resected within several clinically relevant sets, including the SOZ as a baseline for comparison, was associated with postsurgical outcome. For each patient, four categories of channels were investigated:

1. SOZ channels (SOZ),
2. The HAT-derived epileptogenic zone (HAT^EZ^),
3. SOZ channels that were also part of the HAT^EZ^ (HAT^EZ^ ∩ SOZ),
4. Channels that were either in the SOZ or part of the HAT^EZ^ (HAT^EZ^ ∪ SOZ).

For each category, we computed, for each patient, the mean proportion of channels in that set that were resected and stratified these values by surgical outcome. Prior work has not consistently found the proportion of the SOZ resected to predict surgical outcome^6,7^, so this category was included as a baseline for comparison. The remaining categories examined whether this changed with the addition of HAT outputs. For each category, we treated the patient-level proportion of resected channels as a continuous predictor and surgical outcome (seizure-free vs non-seizure-free) as the binary label, summarising predictive performance using the AUROC. To assess statistical evidence that a higher proportion of relevant channels was associated with seizure freedom, we compared the 12 seizure-free and 23 non-seizure-free patients using one-sided Mann– Whitney U tests, with the alternative hypothesis that seizure-free patients had a higher proportion resected. The Mann–Whitney U test was used because these proportions were bounded between 0 and 1, could be skewed, and often contained tied values, so normality was not assumed. Because this was an exploratory post hoc analysis, the four p-values are reported without multiplicity adjustment and interpreted descriptively. An alpha level of 0.05 was used for all inferential tests.

#### Code availability

The analysis code is available at https://github.com/norrisjamie23/SOZ_localisation_SPES_HAT

#### Data availability

The SPES recordings and electrode metadata analysed in this study are publicly available from OpenNeuro as dataset ds004080, version 1.2.4 (https://doi.org/10.18112/openneuro.ds004080.v1.2.4). The five-year outcome labels used in the exploratory analysis are not part of the public dataset.

## Results

**Table 1** summarises the performance of the three models. The HAT model showed the best overall performance, achieving an AUROC of 0.762, with the highest specificity and sensitivity, as reflected in its Youden’s index of 0.364. The MTE model followed (AUROC = 0.734), while the Baseline model had the lowest performance (AUROC = 0.721).

**Table 1.** Performance in identifying the clinical SOZ for each model architecture. Performance metrics include AUROC, Youden’s index, specificity, and sensitivity. Results are reported as mean values with standard errors in parentheses.

| Model | AUROC | Youden's index | Specificity | Sensitivity |
| --- | --- | --- | --- | --- |
| Baseline | 0.721 (0.035) | 0.309 (0.058) | 0.756 (0.020) | 0.553 (0.060) |
| MTE | 0.734 (0.036) | 0.309 (0.063) | 0.758 (0.023) | 0.550 (0.066) |
| HAT | 0.762 (0.031) | 0.364 (0.059) | 0.774 (0.018) | 0.591 (0.061) |

The HAT improved AUROC over the Baseline by 0.041 (one-sided 95% lower confidence bound 0.009; one-sided sign-flip p = 0.00983; Holm-adjusted p = 0.0197; Cohen’s d_z_ = 0.396; n = 35). The improvement over the MTE model was 0.029 (lower bound 0.003; sign-flip p = 0.0205; Holm-adjusted p = 0.0205; Cohen’s d_z_ = 0.357; n = 35).

Our hyperparameter search generated deeper networks with more cross-channel attention heads for both the HAT and MTE models compared with the Baseline.

Example predictions for the HAT model are shown in **Figure 3**. These correspond to two patients, one for whom the model performed better than average (Patient 1, 75^th^ percentile Youden’s index) and one for whom the model performed poorly (Patient 2, 25^th^ percentile Youden’s index). For Patient 1, the model produced five false positives clustered near the clinical SOZ and a single false negative (B). Of these false positives, one was resected, as was the false negative. For Patient 2, a cluster of highly scoring non-SOZ channels is observed (E), resulting in a threshold that includes many false positives (F). None of these regions were resected, though it is unclear whether any of these regions were epileptogenic.

**Figure 3.**
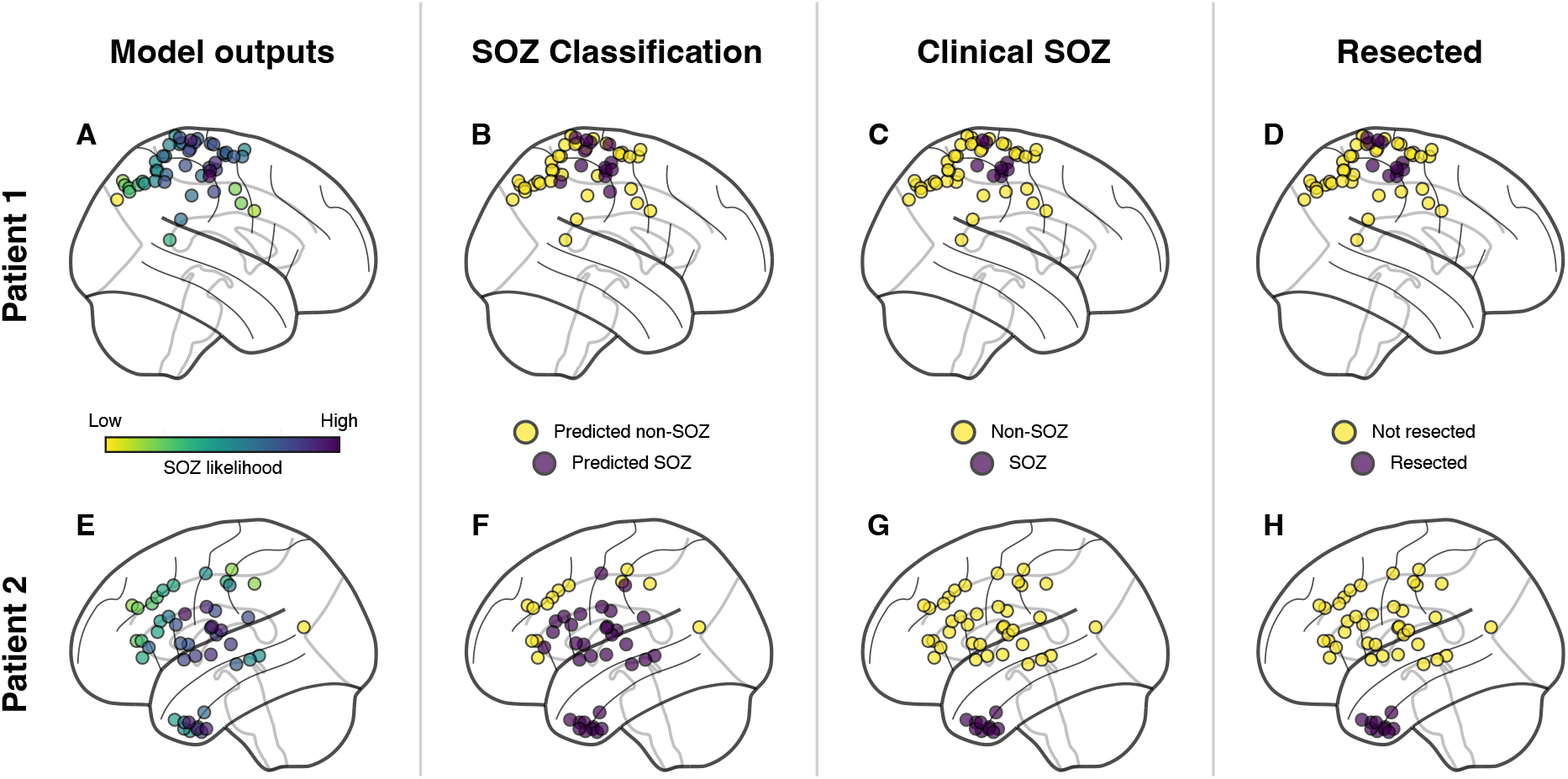
Example predictions and clinical labels for two patients. For Patient 1, the model performed well (**A-D**), and for Patient 2, it performed poorly (**E-H**). Each row displays: **(A, E)** continuous model outputs; **(B, F)** thresholded binary predictions; **(C, G)** clinically identified SOZ; and **(D, H)** resected channels.

### Trial Subset Analysis

Each model was then evaluated using either the first 1 or 5 trials, with performance compared against all available trials (up to 20). As shown in **Figure 4**, degradation was minimal overall. The most notable drop occurred for the Baseline, whose Youden’s index declined from 0.309 with all trials to 0.261 with 1 trial because of reduced sensitivity. Both MTE and HAT remained stable in terms of AUROC and Youden’s index. Full summaries, including for *n* = 2, are provided in Supporting Results S1 and Supplementary Tables S7–S10.

**Figure 4.**
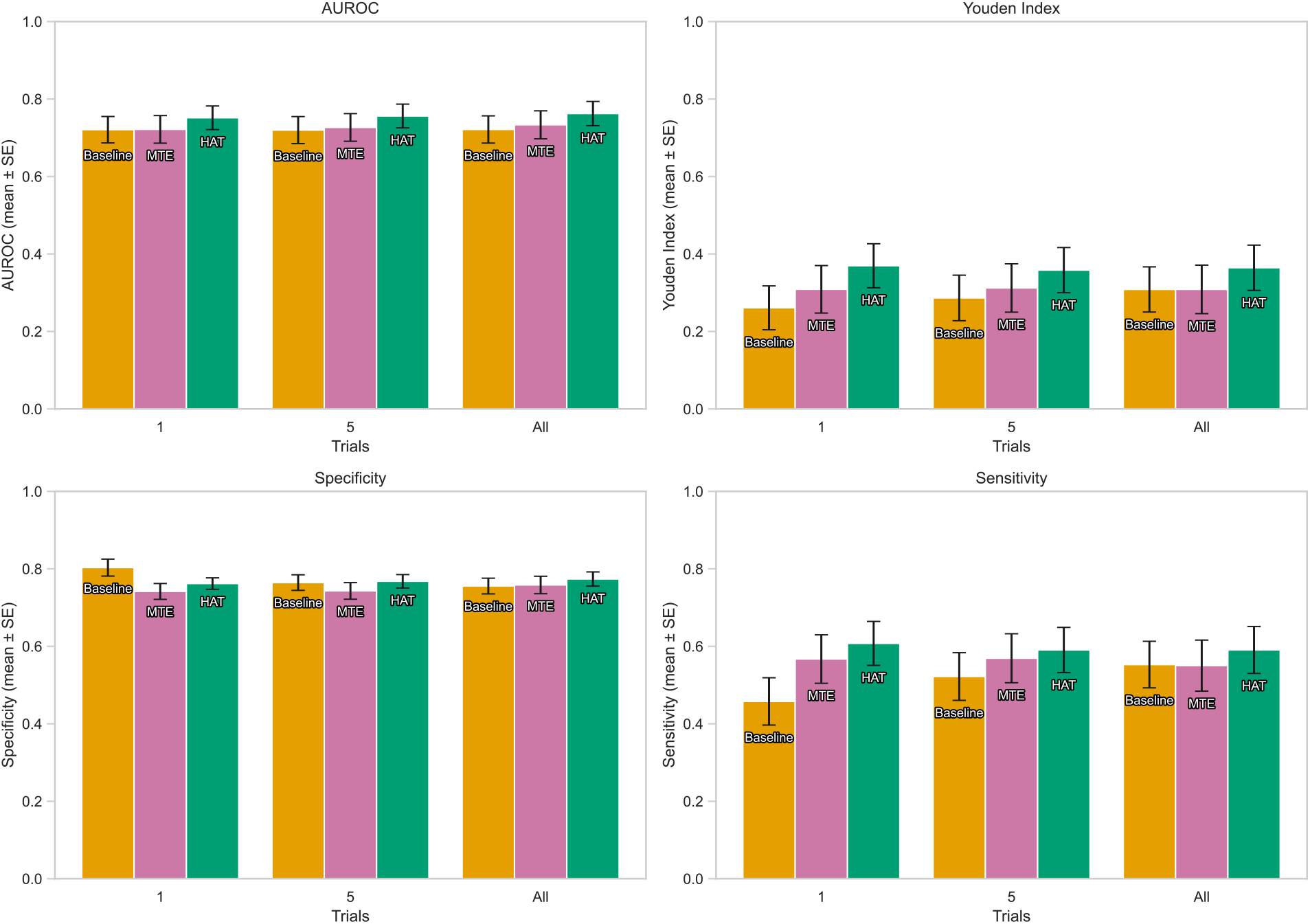
Effect of the number of trials on model performance. The previously trained models were evaluated on the test set using the first n trials only. We show results for n = 1 and n = 5, as well as all available trials (up to a maximum of 20 per site in this dataset). Metrics include AUROC, Youden’s index, specificity, and sensitivity for each model architecture.

### Associations with Surgical Outcome

We evaluated whether the proportion of resected channels in four predefined sets (SOZ, HAT^EZ^, HAT^EZ^ ∩ SOZ, HAT^EZ^ ∪ SOZ) was associated with postsurgical outcome. The results are summarised in **Figure 5**. At the patient level, the AUROC and corresponding one-sided p-values were 0.569, *p* = 0.250 for SOZ; 0.634, *p* = 0.102 for HAT^EZ^; 0.594, *p* = 0.184 for HAT^EZ^ ∩ SOZ; and 0.558, *p* = 0.295 for HAT^EZ^ ∪ SOZ. None of these comparisons reached the 5% significance threshold, providing no statistical evidence that seizure-free patients had a higher proportion resected. Among seizure-free patients, the mean proportion of HAT^EZ^ channels resected was 41.0%, showing that seizure freedom was observed despite most HAT^EZ^ channels not being resected.

**Figure 5.**
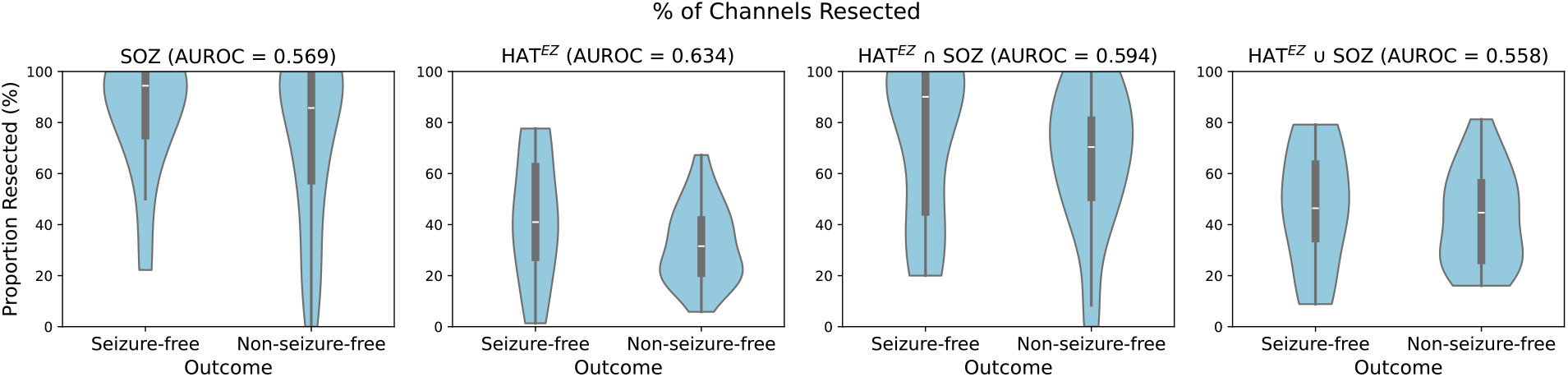
Proportion of channels resected in different categories for seizure-free and non-seizure-free patients. (SOZ, HAT^EZ^, HAT^EZ^ ∩ SOZ, HAT^EZ^ ∪ SOZ). Violin plots show percentage distributions with overlaid quartiles. AUROC values for outcome prediction are shown above each plot; none were statistically significant.

## Discussion

We show that an interleaved Hierarchical Attention Transformer (HAT) with both cross-trial and cross-channel attention improves concordance with the clinical SOZ over a previously published approach that processes trial-averaged SPES responses (the Baseline) and one that embeds individual trials prior to trial-averaging (the MTE). The method generates HAT^EZ^ channel scores from SPES data without requiring observation of spontaneous seizures. It also accommodates heterogeneous electrode configurations and variable numbers of trials. Across repeated five-fold cross-validation, the improvement in performance of the HAT was statistically significant. Averaging SPES responses across trials is convenient for visual review but can suppress features that occur intermittently or vary in latency. By interleaving cross-trial and cross-channel attention, the HAT aggregates information from all available trials during representation learning, rather than imposing a fixed mean prior to the attention mechanism. The HAT improved AUROC by 0.029 over MTE, indicating that cross-trial attention provided only a small additional improvement over mean pooling of trial embeddings. HAT performance changed little when inference was restricted to the first trial at each stimulation site. This suggests that, for this task and dataset, models trained using all available trials may still perform well when fewer trials are available at inference. Whether this could shorten SPES acquisition requires prospective evaluation.

### Clinical utility

Any clinical application warrants careful consideration. Under our thresholding methodology, the HAT showed limited sensitivity, missing 40.9% of SOZ channels. As the SOZ is only a proxy for the epileptogenic zone, lower sensitivity against this label does not necessarily imply poor identification of epileptogenic tissue. However, even amongst the seizure-free subcohort, most HAT^EZ^ channels were not resected. Acting directly on the model’s thresholded outputs without clinical judgement would therefore risk unnecessary removal of large volumes of cortical tissue. The present results do not establish a clinical role for the model. Continuous per-channel scores, such as those shown in **Figure 3A** and **Figure 3E**, may be more informative for future evaluation than binary classifications.

### Associations with Surgical Outcome

When we assessed the association between model outputs and surgical outcome, using the HAT^EZ^ led to the highest mean AUROC, but no approach showed a statistically significant association with outcome. Combining the HAT^EZ^ with the SOZ by intersection or union offered no evidence of added value, and nor did the SOZ alone. Previous studies have reported mixed findings on the relationship between resection of seizure onset contacts and outcome. Cuello Oderiz *et al*.^8^ reported that resection of the seizure onset zone identified during stimulation was associated with surgical outcome. In contrast, other cohorts found no association between the proportion of the clinically defined SOZ resected and seizure freedom^6,7^. The lack of statistical significance in the present study is therefore not unexpected and may reflect limitations of the training labels. The models were trained to identify the clinically defined SOZ, which is only an approximation to the true epileptogenic zone, so any association between the HAT^EZ^ and seizure freedom is indirect and likely weakened by the mismatch between the training target and the clinical endpoint of interest.

### Limitations

Several broader limitations of the present work warrant consideration. Generalisability is restricted by the use of a single-centre cohort of 35 patients with no external validation; a model trained on one centre’s implantation practices, stimulation protocol, and patient population may not transfer to others. Children and adults were analysed together, and the cohort was too small for a reliable age-stratified evaluation. Our recordings used low-frequency (0.2 Hz) electrocorticography (ECoG) stimulation delivered primarily through grid and strip electrodes. In contrast, studies linking trial-to-trial variability and delayed responses to epileptogenicity have often used higher-frequency stimulation (for example, 1 Hz) and stereo-EEG depth electrodes^16,17^. Differences in montage and frequency may affect both the underlying physiology and the observed variability, so our findings may not apply unchanged to other centres or modalities. Because signals were low-pass filtered at 150 Hz, post-stimulation activity above this range, including high-frequency oscillations, was not analysed. Finally, beyond the Euclidean distance supplied for each stimulation–recording pair, we did not encode cortical-surface relationships, anatomical adjacency or regional differences in intrinsic cortical excitability, and we relied on a single modality, whereas epilepsy surgery decisions are inherently multimodal. These limitations reinforce that the model’s predictions should not be used in isolation to dictate resection strategy.

### Future directions

Addressing these limitations will require several methodological improvements. A clinically useful model will need training and evaluation on a larger, multicentre cohort spanning diverse stimulation parameters, implantation strategies (grids/strips and stereo-EEG), and patient populations, with external validation on held-out centres. Reducing label noise is also a priority. Rather than relying on the clinically defined SOZ across the whole cohort, outcome-aligned supervision can be used: in seizure-free patients, channels that were SOZ and resected are treated as positive (epileptogenic), whereas channels that were non-SOZ and not resected are treated as negative (non-epileptogenic). Rather than treating all channels with equal importance, training could prioritise these higher-confidence positives and negatives, with other channels (e.g., from non-seizure-free patients or where the SOZ and resection did not align) down-weighted or excluded from the training data. As a result, model predictions may align better with epileptogenicity.

Regional differences in intrinsic excitability can be handled within the current framework by using patient-specific thresholds that vary by region, rather than a single threshold per patient. With larger and more diverse datasets, these differences could instead be learned during training by incorporating spatial or anatomical embeddings into the cross-channel Transformer to encode cortical location and adjacency. Where available, distance along the cortical surface could be used in place of Euclidean distance to better reflect anatomical proximity. Interpretability is another priority. Integrated Gradients^24^ estimates how different parts of the input contribute to a prediction. Applied here, it could indicate which trials, time windows and stimulation sites most influenced the model output. This could help determine whether the model relies on plausible electrophysiological features, such as delayed responses, rather than artefact. Finally, integrating routinely available modalities (structural and functional imaging, ictal and interictal EEG, semiology) would improve robustness, reduce centre-specific biases, and better align outputs with real-world surgical decision-making.

## Conclusion

In summary, the interleaved HAT delivered small but statistically significant improvements over trial-averaged approaches by explicitly modelling dependencies across trials. Performance changed little with single-trial inputs at inference, although this analysis did not test whether the stimulation protocol could be shortened. The exploratory outcome analysis did not establish clinical utility, and the model should not be used to direct resection. The absence of a statistically significant association with surgical outcome highlights the limitation of training on clinically defined SOZ labels. Future directions include multicentre training with external validation, training on labels that align more closely with epileptogenicity, explainability measures, and integration with other clinical modalities.

## Acknowledgements

J.N. is supported by a UCL UKRI Centre for Doctoral Training in AI-enabled Healthcare studentship (EP/S021612/1). R.R. is supported by the National Institute of Health and Care Research (NIHR, CL-2023-17-004).

## Author contributions

J.N.: Conceptualisation (lead); methodology (lead); software (lead); validation (lead); formal analysis (lead); investigation (lead); visualisation (lead); preparation of the original draft (lead); review and editing of the manuscript (lead).

R.R.: Conceptualisation (lead); methodology (supporting); visualisation (supporting); preparation of the original draft (supporting); review and editing of the manuscript (supporting); supervision (lead).

D.v.B.: Conceptualisation (supporting); data curation (lead); methodology (supporting); review and editing of the manuscript (supporting).

A.C.: Methodology (supporting); review and editing of the manuscript (supporting).

G.C.: Review and editing of the manuscript (supporting); supervision (supporting).

M.T.: Methodology (supporting); review and editing of the manuscript (supporting); supervision (supporting).

K.F.: Review and editing of the manuscript (supporting); supervision (supporting).

S.D.W.S.: Methodology (supporting); review and editing of the manuscript (supporting).

