## Supporting Information for "Localising the epileptogenic zone from single-pulse electrical stimulation responses using cross-trial attention"

**Affiliations:**
[1] UCL Institute of Health Informatics, London, UK
[2] Wellcome Centre for Human Neuroimaging, UCL Queen Square Institute of Neurology, London, UK
[3] Department of Neurology and Neurosurgery, UMC Utrecht Brain Center, University Medical Center Utrecht, Utrecht, The Netherlands
[4] Stichting Epilepsie Instellingen Nederland (SEIN), Zwolle, The Netherlands
[5] Department of Neurosurgery, Great Ormond Street Hospital, London, UK
[6] Developmental Neuroscience, UCL Great Ormond Street Institute of Child Health, London, UK
[7] Department of Neurophysiology, Great Ormond Street Hospital, London, UK
[8] Great Ormond Street Hospital for Children, London, UK
[9] UCL Queen Square Institute of Neurology, London, UK
[10] Department of Basic and Clinical Neuroscience, Institute of Psychiatry, Psychology and Neuroscience, King's College London, London, UK

#### Supporting Methods S1: Additional cohort and stimulation details

Patient characteristics and stimulation parameters are summarised in **Table S1** and

**Table S2**.

***Table S1 Patient and SPES characteristics.*** *Ages are reported in ranges of five years. Stimulation sites are unique bipolar pairs in the released events files, with reversed polarities counted as one site. SOZ contacts are contacts identified as belonging to the clinical SOZ in the released electrode metadata.*

| Patient ID | Age range (years) | Sex | Stimulated sites (bipolar pairs) | SOZ contacts | Current |
|---|---|---|---|---|---|
| 7 | 10–14 | F | 95 | 24 | 8 mA (100%) |
| 15 | 15–19 | M | 45 | 8 | 8 mA (100%) |
| 17 | 0–4 | M | 54 | 3 | 8 mA (100%) |
| 21 | 10–14 | F | 42 | 10 | 4 mA (12%), 8 mA (88%) |
| 26 | 5–9 | M | 48 | 9 | 8 mA (100%) |
| 28 | 10–14 | M | 54 | 5 | 8 mA (100%) |

| Patient ID | Age range (years) | Sex | Stimulated sites (bipolar pairs) | SOZ contacts | Current |
|---|---|---|---|---|---|
| 29 | 10–14 | M | 54 | 4 | 4 mA (9%), 8 mA (91%) |
| 31 | 40–44 | F | 44 | 3 | 8 mA (100%) |
| 33 | 15–19 | M | 56 | 3 | 4 mA (18%), 8 mA (82%) |
| 35 | 20–24 | F | 59 | 10 | 8 mA (100%) |
| 36 | 5–9 | M | 51 | 7 | 4 mA (19%), 8 mA (81%) |
| 37 | 25–29 | M | 49 | 5 | 4 mA (17%), 8 mA (83%) |
| 38 | 20–24 | M | 54 | 7 | 4 mA (22%), 8 mA (78%) |
| 39 | 10–14 | M | 44 | 4 | 8 mA (100%) |
| 40 | 40–44 | F | 49 | 2 | 8 mA (100%) |
| 41 | 30–34 | M | 70 | 5 | 8 mA (100%) |
| 42 | 15–19 | M | 70 | 20 | 8 mA (100%) |
| 44 | 30–34 | F | 48 | 16 | 8 mA (100%) |
| 45 | 20–24 | M | 47 | 7 | 8 mA (100%) |
| 46 | 35–39 | F | 54 | 10 | 8 mA (100%) |
| 48 | 15–19 | F | 58 | 9 | 4 mA (9%), 8 mA (91%) |
| 49 | 10–14 | M | 41 | 1 | 8 mA (100%) |
| 51 | 15–19 | M | 47 | 8 | 8 mA (100%) |
| 52 | 40–44 | M | 51 | 5 | 8 mA (100%) |
| 53 | 25–29 | F | 46 | 3 | 4 mA (11%), 8 mA (89%) |
| 55 | 10–14 | M | 56 | 9 | 8 mA (100%) |
| 57 | 30–34 | F | 51 | 3 | 4 mA (12%), 8 mA (88%) |
| 58 | 15–19 | F | 89 | 37 | 8 mA (100%) |
| 59 | 50–54 | F | 56 | 15 | 4 mA (41%), 8 mA (59%) |
| 60 | 15–19 | M | 69 | 18 | 8 mA (100%) |
| 61 | 5–9 | F | 55 | 19 | 8 mA (100%) |
| 62 | 5–9 | M | 63 | 11 | 4 mA (30%), 8 mA (70%) |
| 63 | 30–34 | F | 66 | 13 | 4 mA (3%), 8 mA (97%) |
| 65 | 15–19 | F | 46 | 18 | 8 mA (100%) |
| 69 | 50–54 | F | 83 | 11 | 8 mA (100%) |

***Table S2 Implant and stimulation protocol***.

| Characteristic | Value |
|---|---|
| Implanted contacts listed in the released metadata | 2,474 in total; median 64 per patient (range 48–112) |
| Recording sites used in the analysis | 80.5% grid and 19.5% strip contacts |
| Subdural electrode contact surface | 4.2 $mm^2$ |
| Subdural intercontact distance | 10 mm |
| Recording sampling rate | 2048 Hz |
| Stimulation arrangement | Bipolar stimulation between two adjacent contacts |

| Characteristic | Value |
|---|---|
| Stimulation waveform | Monophasic pulse, 1 ms duration |
| Stimulation frequency | 0.2 Hz |
| Stimulation current | 4 or 8 mA |
| Repeated stimulation | The acquisition protocol used ten pulses per stimulation sequence. Trials affected by the prespecified seizure or artefact exclusions were removed before analysis, so the number retained could vary. |

#### Supporting Methods S2: Cross-validation procedure

The repeated patient-held-out cross-validation procedure is illustrated in **Figure S1**.

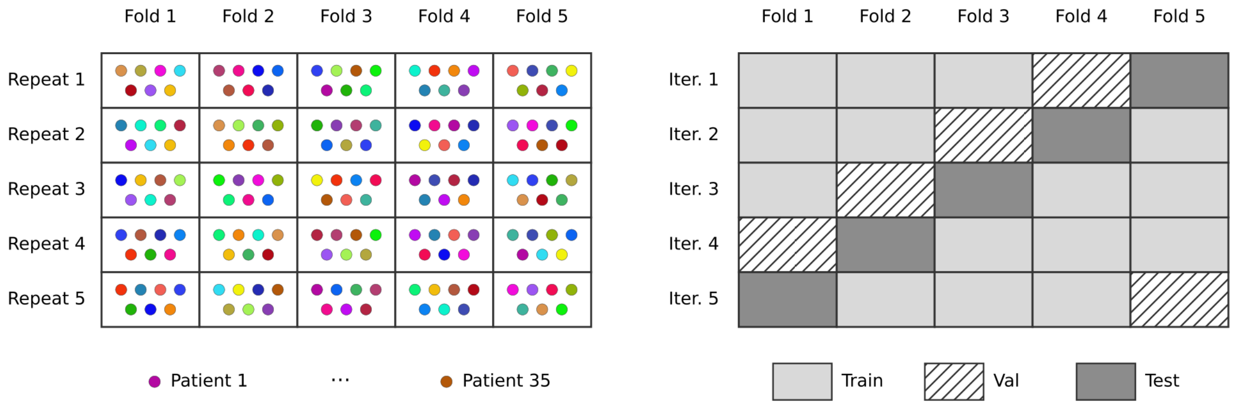


***Figure S1 Repeated patient-held-out five-fold cross-validation. (Left)*** *Between repeats, patients were randomly reassigned to one of five folds, so fold membership changed across repeats. Each coloured point represents one of the 35 patients, with seven patients in each fold.* ***(Right)*** *Within each repeat, five iterations were run. In each iteration, one fold was used for testing, one for validation, and the remaining three for training. Each fold was used once for testing within a repeat.*

#### Supporting Methods S3: Thresholding

As each model produces a continuous output, a threshold is required to compute the specificity, sensitivity, and Youden's index. Given the heterogeneity of model outputs across patients, we used patient-specific thresholds. We used the patient-specific thresholding procedure described in the main Methods, which does not require knowledge of the test patient's SOZ. In brief, for each patient in the validation set we collated model predictions $Y$ (per-channel SOZ likelihoods) and identified the threshold $thresh$ that maximised that patient's Youden's index. We then expressed this threshold as $thresh = median(Y) + n \times MAD(Y)$, where $MAD(Y)$ is the median absolute deviation of the predictions, and solved for the corresponding scalar $n$ for that patient. The median $n$ across validation patients was subsequently used to compute patient-specific thresholds for each patient in the test set. A channel with a score at or above the patient-specific threshold was designated model positive; a score below the threshold was designated model negative. AUROC was calculated from the continuous scores and therefore did not depend on this threshold.

### Supporting Methods S4: Statistical tests

The five held-out AUROC scores for each patient and model were averaged before resampling, giving 35 paired patient-level differences for each comparison. In each of 10,000 bootstrap iterations, 35 differences were sampled with replacement and their mean was calculated. The fifth percentile of the resulting distribution was used as the one-sided 95% lower confidence bound. Repeat-level scores were not resampled separately.

For the sign-flip tests, the sign of each patient-level difference was independently reversed under the null hypothesis. The one-sided p-value was the proportion of permuted mean differences that were at least as large as the observed mean difference. Cohen's $d_z$ was calculated as the mean paired difference divided by the standard deviation of the paired differences.

For descriptive summaries, the five repeat scores were first averaged within each patient. The standard error was then calculated as the standard deviation of the 35 patient means divided by the square root of 35.

### Supporting Methods S5: Hyperparameter search

The hyperparameter tuning in *Optuna* involved 30 trials per model, with pruning introduced after the first five trials to reduce computation time. Trials were pruned if their validation AUROC fell below the median of previous trials at the same run and fold. This pruning check was applied from the 10th fold onward (i.e., starting at run 2, fold 5), allowing the search to focus on more promising configurations. The held-out test fold was not used to propose, prune or select hyperparameter configurations.

The search spaces for hyperparameter optimisation were guided by validation scores from initial experiments and are shown alongside the optimal parameters in **Table S3–S5**.

***Table S3 Hyperparameter search space and optimal hyperparameters for the Baseline model.***

| **Parameter** | **Search Space** | **Optimal** |
|---|---|---|
| Learning Rate | $[5 \times 10^{-5}, 5 \times 10^{-4}]$ | $2.8 \times 10^{-4}$ |
| Dropout | [0.20, 0.50] | 0.39 |
| Embedding Dimension | {8, 16, 32} | 16 |
| Number of Transformer Layers | {2, 4} | 2 |
| Number of Attention Heads | {2, 4} | 2 |

***Table S4 Hyperparameter search space and optimal hyperparameters for the MTE model.***

| **Parameter** | **Search Space** | **Optimal** |
|---|---|---|
| Learning Rate | $[5 \times 10^{-5}, 5 \times 10^{-4}]$ | $2.7 \times 10^{-4}$ |
| Dropout | [0.40, 0.70] | 0.60 |
| Embedding Dimension | {8, 16, 32} | 16 |

| | | |
|---|---|---|
| Number of Transformer Layers | {4, 8, 16} | 8 |
| Number of Attention Heads | {4, 8, 16} | 8 |

***Table S5 Hyperparameter search space and optimal hyperparameters for the HAT model.***

| **Parameter** | **Search Space** | **Optimal** |
|---|---|---|
| Learning Rate | $[5 \times 10^{-5}, 5 \times 10^{-4}]$ | $1.8 \times 10^{-4}$ |
| Dropout | [0.40, 0.70] | 0.52 |
| Embedding Dimension | {8, 16, 32} | 16 |
| Number of Transformer Layers | {4, 8, 16} | 8 |
| Number of Cross-Trial Attention Heads | {2, 4, 8} | 2 |

For the MTE and HAT models, early experimentation suggested that expanding the search space to include additional Transformer layers, attention heads, and higher dropout rates would improve performance. However, early experiments indicated that the cross-trial attention layers required fewer heads than the cross-channel layers (**Table S5**).

#### Supporting Methods S6: Sampling and augmentation

Before sampling, the processed dataset contained 2,117 candidate recording sites, of which 296 (14.0%) were within the clinically defined SOZ and 1,821 were outside it. Within each training partition, sites were sampled with replacement using a weighted random sampler. The weight assigned to each site was based on the frequency of its class and the number of candidate sites contributed by that patient, giving greater weight to SOZ sites and to patients with fewer sites. Binary cross-entropy loss also used a positive-class weight equal to the ratio of non-SOZ to SOZ sites in the training fold. Validation and test sets were not resampled and retained their natural class distributions.

During training, each forward pass masked a randomly selected integer percentage, from 0% to 74%, of the valid observations, where each observation comprised one trial at one stimulation site. Masked observations did not contribute to attention or pooling. Validation and test inputs were not randomly masked. The fixed trial-subset analysis was applied only during inference.

#### Supporting Methods S7: Training times

We report model training durations in **Table S6**. For each method, 30 Optuna trials were run and the table reports the training time for the selected trial. Each selected trial consisted of 25 model runs (5 repeats of 5-fold cross-validation). Inference time for each patient is negligible, taking less than 1 second.

***Table S6 Training time for the chosen trial and the average training time per model within the chosen trial.***

| **Model** | **Total Training Time** | **Per-model Training Time** |
|---|---|---|
| Baseline | 39 minutes | 1 minute 34 seconds |
| MTE | 1 hour 7 minutes | 2 minutes 40 seconds |
| HAT | 2 hours 13 minutes | 5 minutes 20 seconds |

#### Supporting Results S1: Trial subset analysis

**Table S7–S10** present the performance metrics across different trial subsets for each model.

***Table S7 AUROC for each trial subset. Results are reported as mean values with standard errors in parentheses.***

| **Model Type** | **1 Trial** | **2 Trials** | **5 Trials** | **All Trials** |
|---|---|---|---|---|
| Baseline | 0.721 (0.034) | 0.715 (0.035) | 0.720 (0.035) | 0.721 (0.035) |
| MTE | 0.722 (0.036) | 0.721 (0.035) | 0.727 (0.036) | 0.733 (0.036) |
| HAT | 0.752 (0.030) | 0.757 (0.030) | 0.756 (0.031) | 0.762 (0.031) |

***Table S8 Youden's Index for each trial subset. Results are reported as mean values with standard errors in parentheses.***

| **Model Type** | **1 Trial** | **2 Trials** | **5 Trials** | **All Trials** |
|---|---|---|---|---|
| Baseline | 0.261 (0.057) | 0.255 (0.056) | 0.287 (0.059) | 0.309 (0.058) |
| MTE | 0.309 (0.061) | 0.306 (0.061) | 0.312 (0.062) | 0.309 (0.063) |
| HAT | 0.370 (0.057) | 0.371 (0.056) | 0.358 (0.058) | 0.364 (0.058) |

***Table S9 Specificity for each trial subset. Results are reported as mean values with standard errors in parentheses.***

| **Model Type** | **1 Trial** | **2 Trials** | **5 Trials** | **All Trials** |
|---|---|---|---|---|
| Baseline | 0.803 (0.022) | 0.786 (0.022) | 0.765 (0.020) | 0.756 (0.020) |
| MTE | 0.742 (0.020) | 0.735 (0.022) | 0.743 (0.021) | 0.758 (0.023) |
| HAT | 0.762 (0.015) | 0.761 (0.016) | 0.768 (0.017) | 0.774 (0.018) |

***Table S10 Sensitivity for each trial subset. Results are reported as mean values with standard errors in parentheses.***

| **Model Type** | **1 Trial** | **2 Trials** | **5 Trials** | **All Trials** |
|---|---|---|---|---|
| Baseline | 0.458 (0.061) | 0.469 (0.062) | 0.522 (0.062) | 0.553 (0.060) |
| MTE | 0.567 (0.063) | 0.571 (0.061) | 0.569 (0.063) | 0.550 (0.066) |
| HAT | 0.608 (0.057) | 0.610 (0.056) | 0.591 (0.058) | 0.591 (0.061) |
